# A Markov Chain approach for ranking treatments in network meta-analysis

**DOI:** 10.1101/19008722

**Authors:** Anna Chaimani, Raphaël Porcher, Émilie Sbidian, Dimitris Mavridis

## Abstract

When interpreting the relative effects from a network meta-analysis (NMA), researchers are usually aware of the potential limitations that may render the results for some comparisons less useful or meaningless. In the presence of sufficient and appropriate data, some of these limitations (e.g. risk of bias, small-study effects, publication bias) can be taken into account in the statistical analysis. Very often, though, the necessary data for applying these methods are missing and data limitations cannot be formally integrated into ranking. In addition, there are other important characteristics of the treatment comparisons that cannot be addressed within a statistical model but only through qualitative judgements; for example, the relevance of data to the research question, the plausibility of the assumptions, etc. Here, we propose a new measure for treatment ranking called the *Probability of Selecting a Treatment to Recommend* (POST-R). We suggest that the order of treatments should represent the process of considering treatments for selection in clinical practice and we assign to each treatment a probability of being selected. This process can be considered as a Markov chain model that allows the end-users of NMA to select the most appropriate treatments based not only on the NMA results but also to information external to the NMA. In this way, we obtain rankings that can inform decision-making more efficiently as they represent not only the relative effects but also their potential limitations. We illustrate our approach using a NMA comparing treatments for chronic plaque psoriasis and we provide the Stata commands.

## 1 Introduction

Network meta-analysis (NMA) provides the highest possible level of evidence for the development of clinical guidelines and several health care organisations already incorporate NMA findings in their guidance. NMA provides estimates with increased precision and allows estimating the relative effectiveness for interventions that have never been compared head-to-head. Additionally, NMA provides a hierarchy for a specific outcome of all alternative treatments of a comparative effectiveness review as long as they form a connected network. Treatment ranking has gained much attention as well as a lot of criticism over the last years^1–4^. The main criticisms are: a) methods widely used in the literature have focused on the probability of each treatment being the best without taking into account the whole ranking distribution and have produced misleading results^5,6^, b) ranking is a very influential output and interpretation in isolation from relative effects may lead to spurious conclusions^7^ and c) ranking of treatments most often is not interpreted in light of the limitations of the evidence base (such as risk of bias or insufficient evidence).

To date, the most appropriate approaches for treatment ranking in NMA are probably the SUCRA (Surface Under the Cumulative RAnking curves) values^8^ and the more recent P-scores^9^. SUCRA values express *the proportion of effectiveness/safety an intervention achieves relative to a treatment that would be the best with no uncertainty*. P-scores express *the mean extent of certainty that a treatment is better than the other competing treatments*. The two measures are equivalent and differ only in that SUCRAs are obtained using resampling methods while P-scores are derived analytically^9^. Their main advantage is that they account for the variability in treatment ranks by considering not only the magnitude of relative effects but also their uncertainty and overlap of their confidence/credible intervals.

However, SUCRAs and P-scores are limited by the fact that they are based solely on the NMA relative effects and other factors that are often of interest to policy makers in treatment guidelines can only be incorporated in a qualitative way. For instance, if there are reasons to have less confidence in some relative effects than in others, then the usefulness of a conventional treatment ranking becomes questionable. In addition, treatments poorly connected to the rest of the network tend to appear at the top ranks although the evidence favouring such treatments is often insufficient or even biased^10^. Also, safety is a crucial aspect of treatment performance, though very often disregarded in the evidence synthesis setting due to absence of appropriate and adequate data. Last, the cost of the treatments is another important factor when forming clinical guidelines and should be taken into account at the country level.

To address some of these points, Salanti et al.^11^ developed a framework for evaluating the confidence in the treatment ranking from NMA, while Chaimani et al.^12^ suggested a graphical tool depicting the ranking of treatments based on two characteristics. The drawback of the former approach is that a low-confidence ranking would fail to answer its target question (i.e. which are the most appropriate treatments) and efficiently inform medical decision-making. The limitation of the latter approach is that it does not provide a quantitative way to rank the available treatments considering jointly the two characteristics of interest.

In this paper, we suggest the *Probability Of Selecting a Treatment to Recommend* (POST-R) as a new measure to rank treatments after conducting the systematic review and the NMA. Our approach is based on a Markov Chain (MC) model and allows taking into account both the relative effects and the other aforementioned characteristics that may affect treatment choice in practice. MC models have already been employed in several fields for the development of hierarchies and optimization strategies, such as in social networks^13^ and operations research^14^. Here we use a MC approach in which the conditional probability distribution of future states of the treatment selection process depends only on the current state (Markov property); the stationary distribution of this Markov process is regarded as the probabilities of recommending each treatment. The method is Bayesian in the sense that our initial choice of treatment recommendation is informed by the relative effects to result in posterior probabilities of treatment recommendation. The suggested algorithm allows incorporating information other than that of the relative effects through the initial probabilities assigned to each treatment. The rest of the paper is structured as follows: In Section 2, we describe an exemplar NMA used to illustrate our ranking approach. Section 3, after a brief description of P-scores, introduces our ranking method and describes how some key characteristics on top of relative effects can be incorporated in treatment selection. Finally, in Section 4 we use the example dataset to illustrate the method and in Section 5 we discuss the strengths and limitations of our approach.

## 2 Illustrative example

To illustrate the use of our method and compare the results with the conventional ranking results according to SUCRA (or P-score) we use a previously published NMA. Our example is a Cochrane review^15^ comparing the efficacy and safety of 19 different drugs and placebo for patients with moderate to severe psoriasis. The primary outcomes were the events of achieving clear or almost clear skin (i.e. PASI 90) for efficacy and serious adverse events (SAE) for safety.

The authors report in their results for efficacy *“…ixekizumab was the best treatment at drug level (versus placebo: RR 32*.*45, 95% CI 23*.*61 to 44*.*60; SUCRA=94*.*3; high-certainty evidence), followed by secukinumab (versus placebo: RR 26*.*55, 95% CI 20*.*32 to 34*.*69; SUCRA=86*.*5; high-certainty of evidence), brodalumab (versus placebo: RR 25*.*45, 95% CI 18*.*74 to 34*.*57; SUCRA = 84*.*3; moderate-certainty evidence), guselkumab (versus placebo: RR 21*.*03, 95% CI 14*.*56 to 30*.*38; SUCRA = 77; moderate-certainty evidence)…”*; and for safety *“…methotrexate was associated with the best safety profile at drug level in terms of serious adverse events (versus placebo: RR 0*.*23, 95% CI 0*.*05 to 0*.*99; SUCRA = 90*.*7; moderate-certainty evidence), followed by ciclosporin (versus placebo: RR 0*.*23, 95% CI 0*.*01 to 5*.*10; SUCRA = 78*.*2; very low-certainty evidence) […] Nevertheless, analyses on serious adverse events were based on a very low number of events and were reduced to the short time frame of the trials”*^15^. These findings imply that a) the confidence (or certainty) of the evidence is not consistent across drugs and b) the authors are somewhat concerned about the safety results due to rare events and short follow-up of the trials. However, the ranking based on SUCRA provided in the published paper does not account for these factors.

In addition, considering the SUCRAs for both efficacy and safety the original review suggests that Certolizumab, a treatment connected rather weakly to the rest of the network, achieves the better compromise between the two outcomes (Figure 1). Due to the scarcity of the evidence for that treatment, though, several clinicians might be sceptic about this finding. In our application we show how treatments are ranked using the POST-R measure and compare results from each approach.

**Figure 1.**
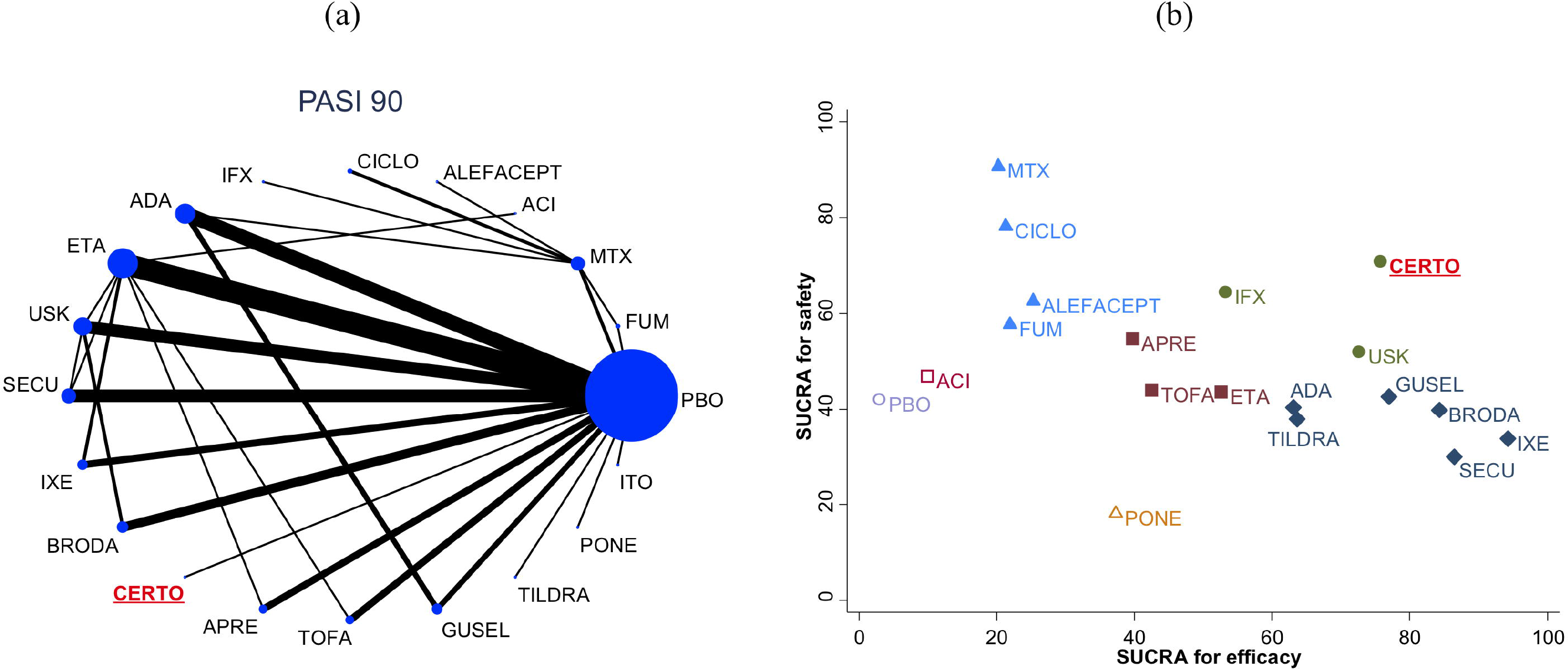
Data and ranking results of the psoriasis network. Panel (a) shows the network diagram for efficacy (PASI 90) and panel (b) the treatment ranking based on P-scores for efficacy and safety jointly.

## 3 Methods

### 3.1 Conventional ranking in NMA

Several different approaches have been used in the literature to rank competing interventions in a network of trials^6^. Here we describe briefly how P-score^9^ (the analytical equivalent of SUCRA^8^), which is based on both the mean and the variance of the NMA relative effects, is computed.

Consider a network with *N* treatments (nodes). P-score is based on the probabilities *p*_*i*>*j*_ that treatment *i* produces a better outcome that treatment *j* with *i, j* = 1, …, *N* and *p*_*i*>*j*_ = 0 for *i* = *j*. Ruecker and Schwarzer^9^ showed that these probabilities can be obtained as

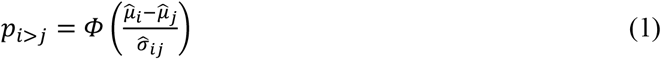

where *Φ* is the cumulative density function of the standard normal distribution, 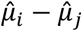 is the estimated difference in the studied outcome between the two treatments and 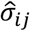 is the estimated standard error of this difference. Once we calculate the *p*_*i*>*j*_ probabilities for every possible pair of treatments (*i, j*) then the P-score for treatment *i* is

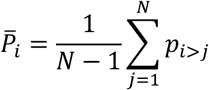

### 3.2 The POST-R measure

#### 3.2.1 Treatment ranking as a discrete stochastic process

We assume a Markov process (*S*) with a countable state space where every treatment (node) *i* = 1, …, *N* within the network under investigation is a state *s*_*i*_. *S* starts at time *t* = 1 by moving between the *N* treatment options with an aim to identify those with the best performance for the main outcome of interest (e.g. efficacy). Each time-point *t* = 1,2,3, … corresponds to a new step in *S*. A movement from *i* to *j* in this setting implies that we were not satisfied with *i* and we select *j* as a potentially more beneficial treatment. Since movements represent the preference between two treatments, they are not uniformly distributed but they are based on prior beliefs and actual data. So, let 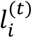 be the probability of selecting treatment *i* at time (i.e. step) *t* and 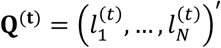. The vector **Q**^(**0**)^ is called the *initial state probability vector* with entries the probabilities 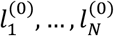 of selecting each treatment *i* at time *t* = 0; hence without looking at the NMA relative effects for the outcome of interest. Different ways for defining **Q**^(**0**)^ are suggested in Section 3.3.

A key aspect of the model is the definition of the *transition probabilities* 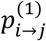 at the beginning of *S*; that corresponds to the probability of selecting treatment *j* at *t* = 2 after having selected treatment *i* when *t* = 1. These probabilities form the component of the MC model that incorporates the NMA relative effects for updating the initial probabilities. We suggest that 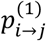’s can be driven by the probabilities *p*_*i*>*j*_ in Equation (1) implying that at *t* = 1 movements between treatments are solely based on the probabilities that each treatment *i* produces a better outcome than *j*. Given that the transition probabilities at every *t* should satisfy 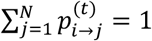, we define

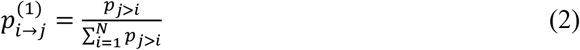

It follows from Equations (1) and (2) that 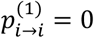 while for any pair 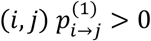. As a result, the transition matrix at the beginning of *S* is

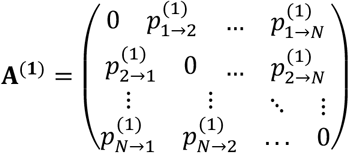

while the transition matrix at each *t* is the product of transition matrices until *t*, 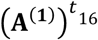. For simplicity, in the rest of the manuscript we suppress the superscript (1) when *t* = 1. Equation (2) implies that **A**^*t*^ corresponds to an irreducible (i.e. 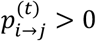 for some *t* > 0) and ergodic (since for all states 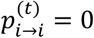 for *t* = 1,2,3, …) MC^16^. When the MC is ergodic it has a unique stationary distribution *π* to which it converges for large values of *t*, namely 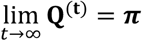. It has been shown that

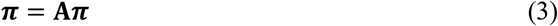

and thus ***π*** is an eigenvector of **A** with eigenvalue 1^17^. For a treatment network this stationary probability distribution of the MC contains the probabilities 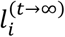 of selecting each treatment after many steps 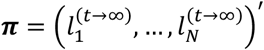 and offers an intuitive way to rank the treatments of a NMA.

According to Equation (3) treatment ranking could be easily obtained as an eigenvector (with eigenvalue 1) of matrix **A**; however, it is informed only by the relative effects (via the transition probabilities in **A**) and therefore it shares the same limitation with the conventional ranking approaches discussed earlier.

#### 3.2.2 Incorporating the initial probability distribution in ranking

To build our model, we employ a special case of a MC previously used for the development of the PageRank algorithm that ranks websites by Google search results^13^. The difference between Google’s model and other MC models is that the former considers that there is always a probability of starting again the stochastic process according to the initial probability distribution. So, let *z* ∈ (0,1) be the probability that the treatment selection process continues with updating at each step the probability distribution according to **A** and (1 − *z*) the probability that the process starts again from **Q**^(**0**)^. It should be noted that such a scenario reflects situations where the NMA fails to adequately update the existing evidence (e.g. due to few data) and conclusions are based to some extent on previous knowledge or other characteristics (such as data quality). More discussion on the definition of *z* is presented in Section 3.4.

This MC configuration leads to the modified transition probabilities

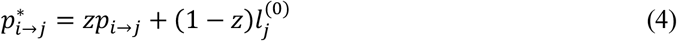

and the modified transition matrix

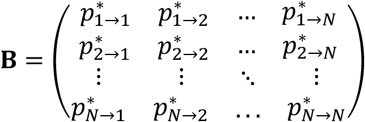

with 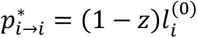.

Given that 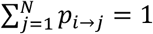 and 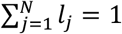 it holds that 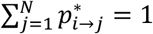. We denote the unique stationary probability distribution (always existing for *z* ∈ (0,1)) of the transition matrix **B** with the vector 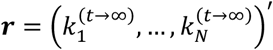. Note that this matrix can be written also in the form:

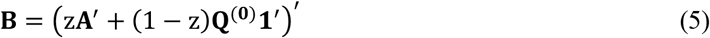

where **1** is a *N* × 1 vector with all elements equal to 1^18^. It follows from ergodic theory for the new MC (*S*^*^) that *S*^*^ has a unique stationary distribution ***r*** with ***r*** = **B*r*** and thus the probability distribution ***r*** is an eigenvector of **B** with eigenvalue 1.

A graphical representation of our MC model for treatment selection is provided in Figure 2. The graph shows a hypothetical example of a network with three alternative treatment options. The process *S* start from **Q**^(**0**)^ and based on existing knowledge about each treatment we move to one of the *A, B, C* with probability 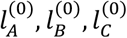 respectively. Let us assume that we first select *A* (at *t* = 1) where we are informed about the NMA relative effects. At the next step we have three options:

**Figure 2.**
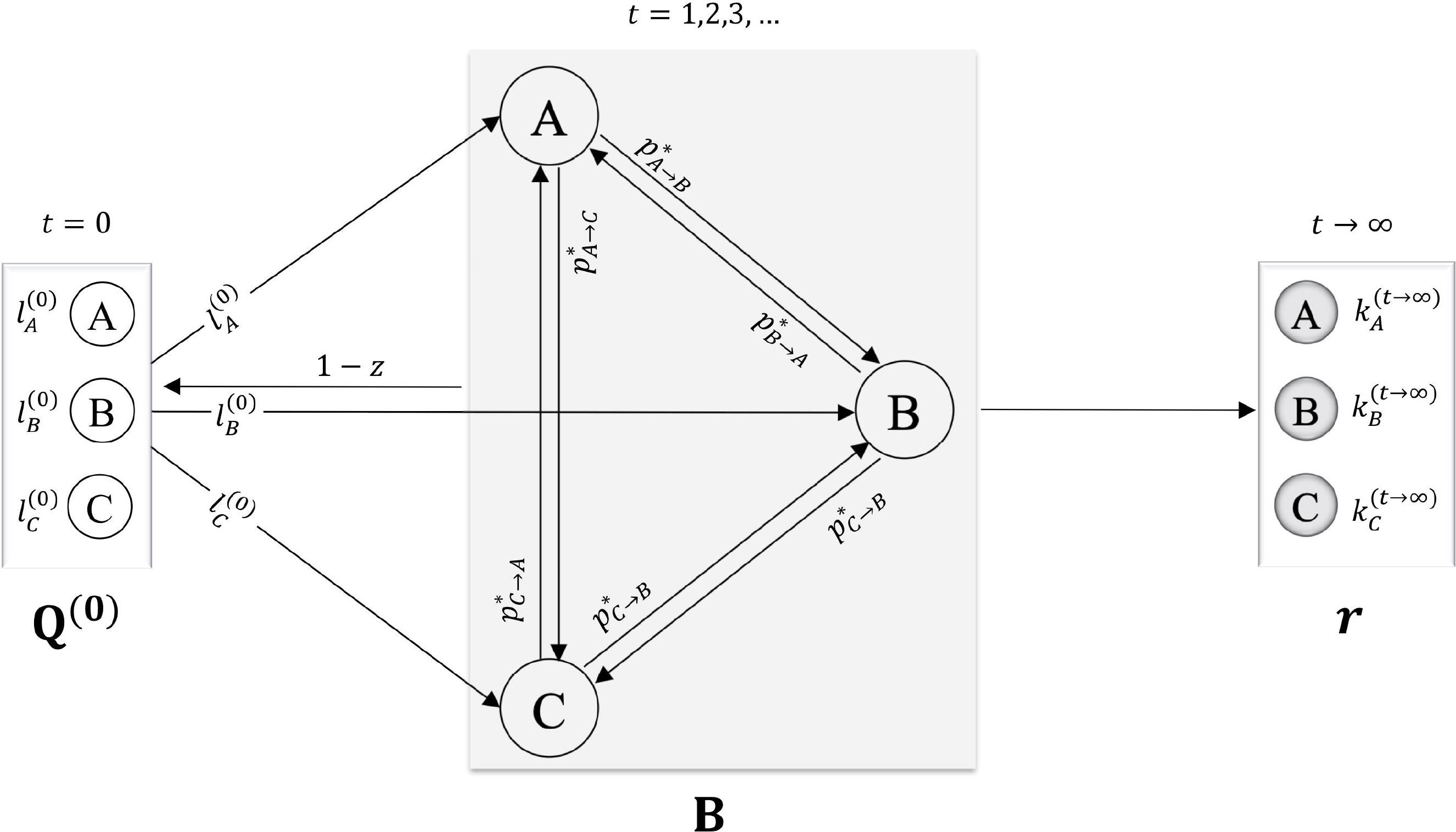
Graphical representation of the Markov Chain model for treatment selection using a hypothetical example of a three-treatment network. Arrows represent the movement from one treatment to the other that happens when a change in the preference between the two treatments occurs.

i. to select *B* with probability 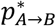 (Equation (4))
ii. to select *C* with probability 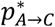
iii. to go back to **Q**^(**0**)^ and start again from *A* with probability 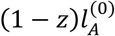

After several steps we will have tried all treatments and will have formed the final selection probabilities 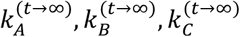.

### 3.3 Defining the Q^(0)^ vector

The initial state probability vector in **Q**^(**0**)^ is the key feature that differentiates our method with the existing ranking approaches (see Section 3.1), and allows incorporating information other than NMA relative effects for treatment recommendation. It is, though, challenging to find a reasonable way to define the initial probabilities 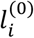. Investigators should bear in mind that each 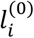 represents the probability of selecting every treatment *i* in clinical practice without knowing the NMA relative effects for the outcome of interest and consequently neither the probabilities *p*_*i*>*j*_. In such a case, treatment choice depends on other characteristic(s) expressed via **Q**^(**0**)^. Here we consider the following characteristics, which we believe are among the most important for forming treatment guidelines:

- Confidence in the evidence: evidence on some of the treatments might be less ‘trustworthy’ than for others. It should be noted that confidence in the evidence from NMA is based on the network structure, precision, the validity of the assumptions, assessment of risk of bias etc., so it is dependent on the NMA setting but independent of the actual values of the relative effects.
- Clinical experience: prior information from clinical practice is important and is not always in agreement with study results as the latter may lack power, have a short follow-up period etc.
- Cost of the treatments: the relative outcomes and costs of available interventions should be assessed, cheaper treatments might be preferable if they yield similar outcomes to slightly more effective, but expensive, ones.
- Treatment safety: efficacy and safety should always be considered jointly when forming recommendations.

It is evident that the definition of the probabilities 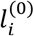 is subjective to some extent and depends on the medical field and research question. In the following sections we suggest ways for defining **Q**^(**0**)^ using the four characteristics mentioned above.

#### 3.3.1 Confidence in the evidence as a criterion for Q^(**0**)^

All recent reporting guidelines for NMA recommend including in the manuscript a table with the confidence (called also quality or certainty in the literature) for every treatment comparison in the network^7,19^. To date, two approaches are available for preparing such a table^11,20^. Here we consider the approach by Salanti et al.^11^ and the recent updates implemented in the online tool CINeMA (available from http://cinema.ispm.ch/)^21,22^. Note that such an evaluation typically is conducted after completing the systematic review and the NMA and not at the beginning of the process. However, it still can be used to form ‘prior’ information on treatment ranks.

We focus on the final report of the full evaluation where there are four possible levels of confidence assigned at each comparison: high, moderate, low, and very low. We translate these levels into probabilities that the evidence is trustworthy. Reasonable values might be *P*(*trust*) = 0.9, 0.7, 0.3 and 0.1 respectively. Then, we can obtain the following confidence matrix

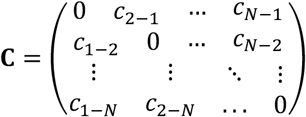

where *c*_*i*−*j*_ is the probability of ‘trusting’ the use of treatment *i* more than treatment *j*. We define *c*_*i*−*j*_ as follows:

- If the mean relative effect between *i* and *j* favours *i*, then *c*_*i*−*j*_ = *P*(*trust*)_*i*−*j*_ and *c*_*j*−*i*_ = 0
- If the mean relative effect between *i* and *j* favours *j*, then *c*_*j*−*i*_ = *P*(*trust*)_*i*−*j*_ (note that *P*(*trust*)_*i*−*j*_ = *P*(*trust*)_*j*−*i*_) and *c*_*i*−*j*_ = 0
- If the mean relative effect between *i* and *j* does not favour any treatment, then *c*_*i*−*j*_ = *P*(*trust*)_*i*−*j*_/2 and *c*_*j*−*i*_ = *P*(*trust*)_*i*−*j*_/2.

It is important to note that what relative effect would lead to favour one treatment over the other depends on the clinical setting. For example, the investigators might define a priori the value of a clinically important effect to be used (such as an estimated risk ratio of 1.1, 1.2, etc.). Using the columns of **C** we can obtain an estimate of the average confidence for using each treatment *i* as

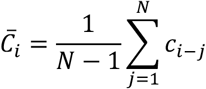

which is the analogous to the P-score 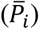. Since **Q**^(**0**)^ is a probability vector, its elements should add up to 1. Therefore, we define

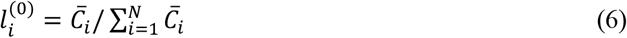

This is the probability to select treatment *i* among all treatments in the network given the matrix **C**.

#### 3.3.2 Clinical experience as a criterion for Q^(0)^

Several methods have been suggested in the literature for expert opinion elicitation on probability distributions^23^. Possibly one of the simplest approaches that can be easily understood by clinicians is the so-called *roulette method* which has been implemented also in the MATCH tool (http://optics.eee.nottingham.ac.uk/match/uncertainty.php)^24^. Briefly, we adapted the method to our setting as follows: We provide to experts a grid with *N* equally sized bins each corresponding to a treatment in the network. Subsequently, we ask from the experts to allocate 100 chips (representing 100 patients) to the *N* bins based on their experience about the performance of the treatments in terms of a particular characteristic; for instance, by considering their safety alone. Then, the initial probability 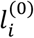 of selecting each treatment *i* is the proportion of chips allocated to the respective bin. Inevitably, expert opinions are subjective to some degree but asking several experts may mitigate this subjectivity. A simple (although not perfect) way, for instance, to combine opinions from 2 or more experts is to use as 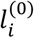 the weighted average of the expert-specific probabilities with weights the years of experience.

#### 3.3.3 Costs a criterion for Q^(0)^

Consider that *T* is the most expensive treatment in the market among the *N* options. We define

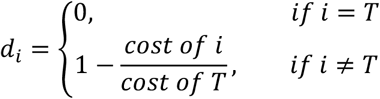

This means that the probability of selecting treatment *i* based on its cost is related to the percentage of the cost of *T* that *i* has. Then, the probabilities 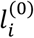 are obtained by Equation (6), replacing 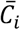 by

#### 3.3.4 Safety a criterion for Q^(0)^

Although we focus on treatment safety here, the approach applies equally to any other outcome. The combination of two outcomes in the treatment selection process can be made through a two-stage approach:

- At the first stage we rank the treatments for the ‘less’ important outcome between the two (for example safety if efficacy is considered more important) using the POST-R approach.
- Then, at the second stage the stationary probability distribution *r* obtained for this outcome becomes the initial probability vector **Q**^(**0**)^ in Equation (5) to be combined with A for the overall treatment selection probabilities.

### 3.4 Defining the probability *z*

Considering that a NMA is (or should be) conducted when there is a need for updating and extending the existing evidence, we believe that *z* should generally take values larger than 0.5. We suggest that values between 0.6 and 0.9 might be reasonable. It is not straightforward, though, how to select the optimal *z* value as this might depend also on the clinical setting and the available data. Running a sensitivity analysis on a range of values for *z* could show how robust ranking results are under different scenarios. The value of *z* can also be informed by expert opinion. For example, we can ask several experienced clinicians to rate (e.g. in a scale) the usefulness/necessity of updating the evidence and clinical practice using the NMA relative effects. Note that *z* cannot takes values 0 or 1; this is a necessary condition for reassuring that an eigenvector of matrix B with eigenvalue 1 exists^25^.

## 4 Application to psoriasis network

### 4.1 Conventional treatment ranking

Erreur ! Source du renvoi introuvable. shows the P-score percentages and the respective ranks for all treatments for the two primary outcomes (PASI 90 and SAE). These were obtained using the *network*^26^ and *network graph*^27^ packages in Stata. The dataset and the Stata script used to obtain the results can be found in the Supplementary files. The league table of the relative effects and the respective league table with the probabilities *p*_*i*>*j*_ are available in the Appendix (Appendix Table 1 **Appendix** Table ***2*** respectively).

### 4.2 Treatment ranking combining efficacy and confidence in the evidence

Using the CINeMA application, we obtained the domain-specific judgements for each comparison in the network and then we summarized them into an overall confidence for every comparison (Appendix Table 3). The last column of the table presents the probability *P*(*trust*) for each comparison. We considered overall in CINeMA a risk ratio of 2 as a clinically important relative effect between two treatments to justify selection of one treatment over the other. This clinically important effect was used for the evaluation of imprecision, heterogeneity and incoherence (more details available in CINeMA’s documentation page http://cinema.ispm.ch/#doc) as well as for the definition of probabilities *c*_*i*−*j*_ (see Section 3.3.1). The matrix C is available in the Appendix (Appendix Table 3), while the probabilities 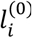 (Table 1) were obtained using Equation (6). Interestingly, ixekizumab that appears to be the most efficacious treatment for PASI 90 is also at the top of the ranking with respect to the confidence of the evidence followed by guselkumab (Table 1). We considered that *z* = 0.75 is a good compromise between the two sources of information (i.e. relative effects for efficacy and confidence of the evidence for efficacy). It seems that the incorporation of the confidence in the evidence resulted in small changes in the overall picture of treatment ranking for efficacy (Figure 3). That was expected given that treatments that appeared to perform better with respect to efficacy were also at the top ranks in terms of the confidence in the evidence (Table 1).

**Table 1.** P-score values and initial probability distributions of treatment selection based on confidence in the evidence, clinical experience and treatment cost. Grey cells correspond to the five top positions in the respective ranking.

| Drug | Efficacy |  | Safety |  | Confidence in the evidence for efficacy |  | Clinical experience for efficacy |  | Clinical experience for safety |  | Safety (relative effects and clinical experience) |  | Treatment cost |  |
| --- | --- | --- | --- | --- | --- | --- | --- | --- | --- | --- | --- | --- | --- | --- |
| | *P*-score | rank | *P*-score | rank | $l_i^{(0)}$ | rank | $l_i^{(0)}$ | rank | $l_i^{(0)}$ | rank | $l_i^{(0)}$ | rank | $l_i^{(0)}$ | rank |
| ACI | 09.9 | 19 | 46.9 | 9 | 0.002 | 18 | 0 | 9 | 0 | 10 | 0.045 | 7 | **0.162** | **1** |
| ADA | 63.1 | 8 | 40.4 | 14 | 0.071 | 6 | **0.100** | **3** | **0.050** | **3** | 0.042 | 10 | 0.05 | 8 |
| ALEFACEPT | 25.3 | 15 | **62.6** | **5** | 0.014 | 17 | 0 | 9 | 0 | 10 | 0.045 | 7 | 0 | 0 |
| APRE | 39.7 | 13 | 54.7 | 7 | 0.027 | 14 | 0 | 9 | 0 | 10 | 0.038 | 13 | **0.106** | **5** |
| BRODA | **84.3** | **3** | 39.8 | 15 | **0.091** | **4** | **0.050** | **4** | **0.050** | **3** | 0.041 | 12 | 0 | 0 |
| CERTO | **75.7** | **5** | **70.9** | **3** | 0.059 | 7 | **0.050** | **4** | **0.050** | **3** | **0.072** | **5** | 0 | 0 |
| CICLO | 21.3 | 17 | **78.2** | **2** | 0.018 | 16 | 0 | 9 | 0 | 10 | **0.075** | **3** | **0.162** | **1** |
| ETA | 52.6 | 11 | 43.6 | 11 | 0.059 | 7 | **0.050** | **4** | **0.150** | **2** | **0.073** | **4** | 0.073 | 6 |
| FUM | 21.9 | 16 | 57.7 | 6 | 0.014 | 17 | 0 | 9 | 0 | 10 | 0.042 | 10 | **0.162** | **1** |
| GUSEL | **77.0** | **4** | 42.6 | 12 | **0.099** | **2** | **0.150** | **2** | **0.050** | **3** | 0.044 | 9 | 0 | 0 |
| IFX | 53.2 | 10 | **64.4** | **4** | 0.049 | 9 | **0.050** | **4** | **0.050** | **3** | 0.059 | 6 | 0.073 | 6 |
| ITO | 56.0 | 9 | - | - | 0.028 | 12 | 0 | 9 | - | - | 0 | 20 | 0 | 0 |
| IXE | **94.3** | **1** | 33.7 | 17 | **0.111** | **1** | **0.050** | **4** | **0.050** | **3** | 0.037 | 14 | 0 | 0 |
| MTX | 20.2 | 18 | **90.7** | **1** | 0.028 | 12 | **0.100** | **3** | **0.150** | **2** | **0.112** | **2** | **0.162** | **1** |
| PBO | 02.9 | 20 | 42.0 | 13 | 0.002 | 18 | 0 | 9 | 0 | 10 | 0.027 | 18 | 0 | 0 |
| PONE | 37.3 | 14 | 18.1 | 19 | 0.027 | 14 | 0 | 9 | 0 | 10 | 0.014 | 19 | 0 | 0 |
| SECU | **86.5** | **2** | 29.9 | 18 | **0.095** | **3** | **0.050** | **4** | **0.050** | **3** | 0.034 | 15 | 0 | 0 |
| TILDRA | 63.6 | 7 | 37.8 | 16 | 0.033 | 11 | 0 | 9 | 0 | 10 | 0.033 | 16 | 0 | 0 |
| TOFA | 42.5 | 12 | 44.0 | 10 | 0.043 | 10 | 0 | 9 | 0 | 10 | 0.030 | 17 | 0 | 0 |
| USK | 72.6 | 6 | 52.0 | 8 | **0.078** | **5** | **0.350** | **1** | **0.350** | **1** | **0.136** | **1** | 0.05 | 8 |

**Figure 3.**
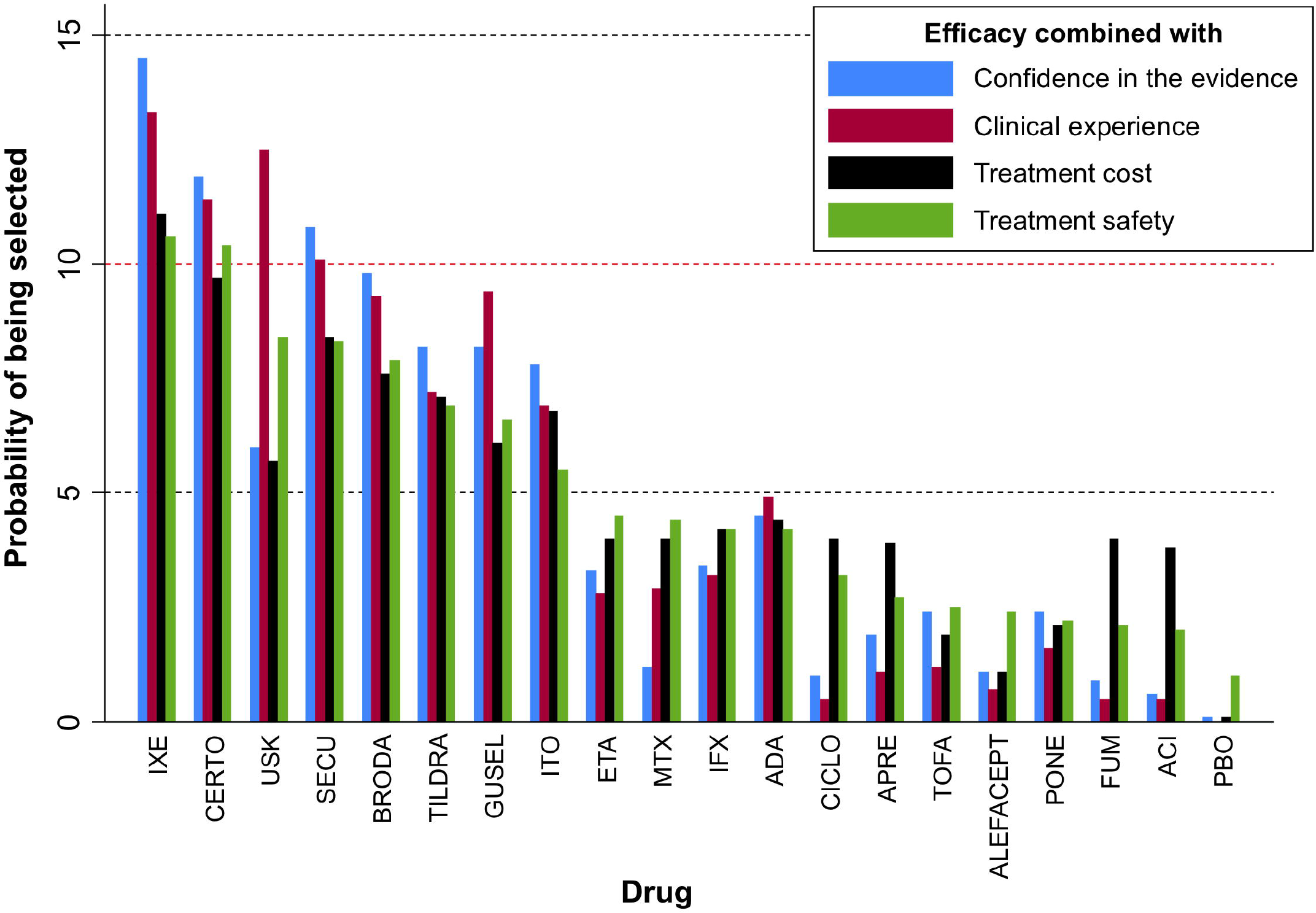
Ranking results of the psoriasis network using the POST-R measure and considering the relative effects for efficacy in the transition probabilities and different characteristics in the initial probability distribution as indicated by the legend. Drugs have been ordered according to the ranking for efficacy and safety (green bars).

### 4.3 Treatment ranking combining efficacy and clinical experience for efficacy

We elicited the clinical experience data on efficacy in terms of treatment selection probabilities using the MATCH tool as Appendix Figure 1(a) shows. Each chip represented 5 hypothetical patients from the population of the original Cochrane review. For illustration purposes in the present study, one dermatologist was asked to allocate 100 patients to the 20 possible treatment options (including placebo) considering how participants are on average allocated to the different drugs in clinical practice. In the presence of prescription data (e.g. from national registries), these could also be used to form some kind of clinical experience data. We used again *z* = 0.75 considering clinical experience for this outcome needs only be a supplementary source of information. Figure 3 implies that the incorporation of clinical experience materially affects the ranks of the treatment. In particular, ustekinumab is favoured and appears at the second position now with more than 12% probability of being selected among all treatments.

### 4.4 Treatment ranking combining efficacy and safety informed by clinical experience

Here we obtained the final *r* vector in two steps:

- First, we combined the clinical experience probabilities for safety with the relative effects for safety to obtain the safety ranking. Considering the concerns expressed previously about the SAE data (see Section 2), we used *z* = 0.7 that reduced slightly more the influence of the relative effects in treatment ranking for safety. The resulted probability vector of this step (*r*_*safety*_), which is shown in Table 1, was then used at the second step as the initial probability distribution **Q**^(**0**)^.
- At the second step, we combined the **Q**^(**0**)^ as defined above with the relative effects for efficacy to rank treatments based on both outcomes using *z* = 0.65.

Again, ustekinumab seems to be the most favoured treatment in comparison with the conventional ranking results based on SUCRAs. However, ixekizumab and certolizumab remained at the top of the hierarchy.

### 4.5 Treatment ranking combining efficacy and cost

In the psoriasis example, the 19 different systemic treatments are very diverse in terms of their cost; therefore we considered treatment cost as an important characteristic to be incorporated in treatment ranking. Specifically, the cost of each treatment per person per year in France was used to form the **Q**^(**0**)^ vector following the approach in Section 3.3.3. Note that treatment cost might be different across countries and therefore our findings might not be directly applicable outside France. Appendix Table 5 shows the average cost of each drug. Secukinumab, ixekizumab and brodalumab are the most expensive treatments, while some of the drugs have not yet been commercialised; for such drugs we considered that they have zero probability of being selected considering their cost only. We used a probability *z* = 0.8 implying that treatment cost should not be considered as a key component of treatment ranking but only for selection between drugs with overall similar performance on other important factors (such as the outcomes of interest). Figure 3 shows that, after incorporating in ranking the cost information, the highly efficacious drugs remained at the top positions.

Overall, according to Figure 3 a group of eight drugs achieved more 5% probability of being selected under all four considerations and one drug more than 10% (ixekizumab). Ustekinumab seems to the drug with most unclear performance as it was favoured when clinical experience was considered but not so much by the actual data. This possibly means that the benefits of this drug need further investigation.

## 5 Discussion

Treatment ranking is a key and potentially very informative output of NMA but inappropriate use and misinterpretation of ranking, which are regularly encountered in published NMAs, have made several researchers being sceptical about its usefulness^1,5,6^. A key point prior to drawing conclusions from ranking results is understanding what the estimated order of the treatments may represent. Currently available approaches provide rankings based either only on the mean of the relative effects (e.g. ranked forest plots) or on the mean and a part of the distribution of the relative effects (e.g. the probability of being best/worst) or at best on the mean and the full distribution of the relative effects (i.e. SUCRAs/P-scores). As a result, these methods produce rankings that are as reliable as the estimated means and/or variances of the relative effects.

In the present paper, we move forward from ranking measures that just provide a summary of the relative effects and introduce a new concept of treatment ranking. A drawback of our approach is that the definition of the vector **Q**^(**0**)^ and the probability *z* is subjective to some degree. It is important that one describes in detail in the protocol of the analysis how (s)he will define these quantities. We can also mitigate this subjectivity by incorporating views from many clinicians with experience in the condition under investigation. In addition, the method can be also applied in Bayesian framework were **Q**^(**0**)^ and *z* can be modelled through distributions allowing for uncertainty in their values. It should be noted that both **Q**^(**0**)^ and *z* are dependent on the characteristics being considered, the medical field and the research question and therefore the criteria for their definition are specific to every NMA. Although such an approach seems to require additional time and resources than current practices in NMA, recent directions in evidence synthesis that encourage living systematic reviews and live cumulative network meta-analyses within the context of a research community are well compatible to our method^28–32^.

An important limitation of our application is that probabilities based on clinical experience were elicited after the publication of the original NMA. Hence, the clinician who provided expertise could have been influenced by the published NMA results. It should be noted that we used a possibly extreme value of the minimally clinical important difference in CINeMA (i.e. risk ratio of 2) which might not apply in clinical practice. Moreover, in the present article we surveyed only one clinician for simplicity; articles aiming to address clinical questions should opt for several experts to inform their POST-R model. In addition, there might be other important characteristics affecting treatment selection that were not considered here. It is important to note that our results are only illustrative of the methods and do aim to draw clinical inferences; the latter would require careful consideration of the definition for **Q**^(**0**)^ and *z* at the beginning of the study. To avoid data-driven decisions and selective reporting of results when using our ranking approach, it is important that a clear and transparent description of the criteria to be used for the definition of **Q**^(**0**)^ and *z* are available in the protocol.

Our approach offers a more intuitive way of thinking treatment ranking as it accommodates the different considerations being made before selecting a treatment in practice. The POST-R approach has been implemented in the ‘sucra’ command of the *network graphs* package^27^ in Stata^33^ which facilitates its use once **Q**^(**0**)^ has been specified. Additional work is necessary to extend the method into a generalised ranking framework that will allow multiple characteristics to be combined simultaneously with the relative effects. A key issue, very often disregarded in the literature, when obtaining the ranking of treatments is to acknowledge that it is always accompanied with a level of uncertainty^2,4,11^. The POST-R approach naturally captures the uncertainty in treatment selection by allowing the end-user of NMA to ‘move’ between the different treatment options (or between groups of treatments) with some probability.

Generally, despite the various and well-founded criticisms, estimation of ranking remains one of the main objectives for several NMAs implying that clinicians and decision-makers are interested in it. Thus, it is necessary to produce NMA summaries that, instead of just acknowledging any data limitations and/or availability of external evidence, they formally integrate these considerations with the results in the league table to make valid recommendations. Such an effort might be less important in dense and coherent networks with many high-quality studies for every treatment comparison. Our method may target primarily NMAs stating *“more well-conducted studies are necessary”* with an aim to assist such reviews drawing more useful conclusions until new studies come up.

## Data Availability

The data are available upon request.

## Appendix Figures

**Appendix Figure 1.**
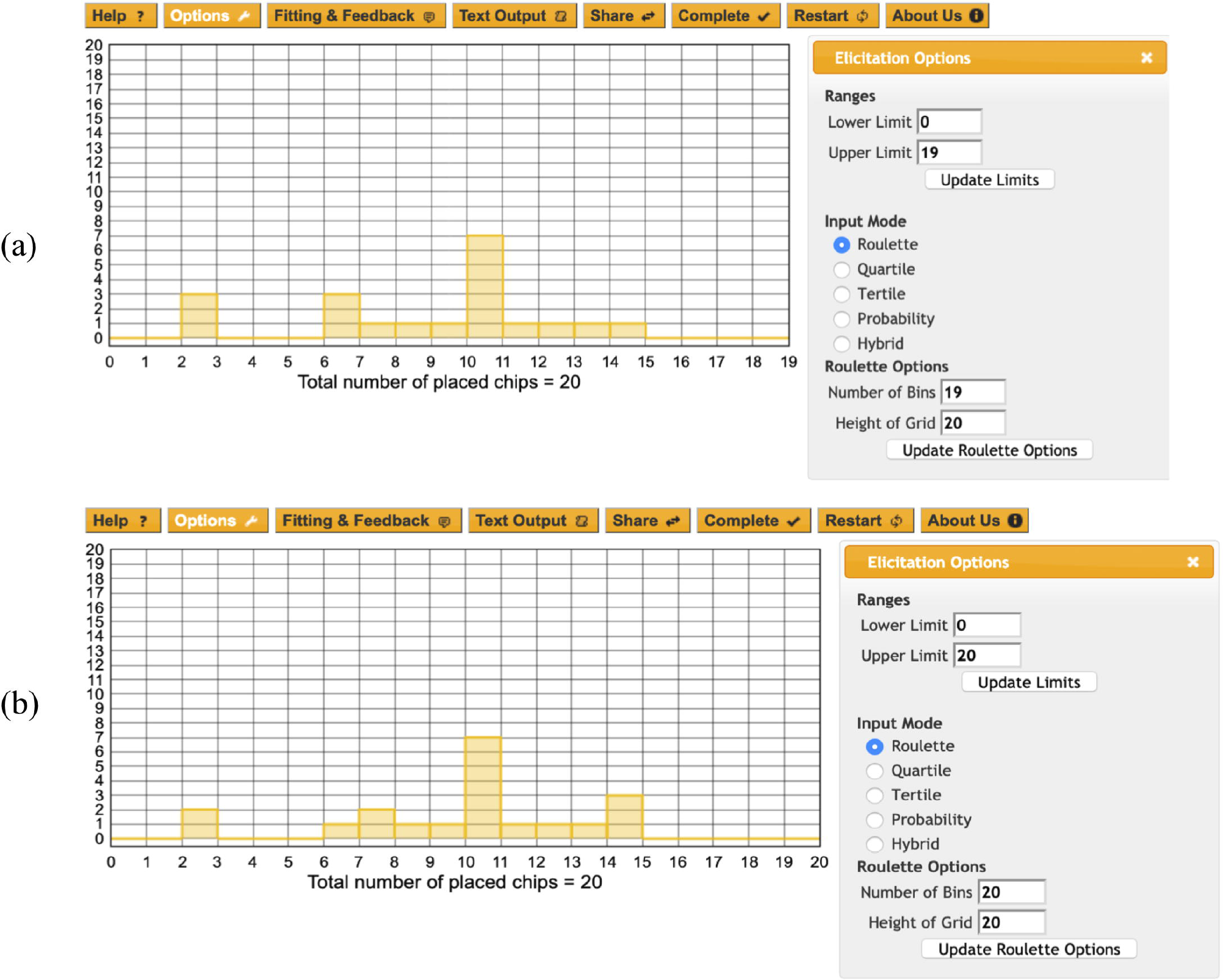
Elicitation of the probabilities of selecting each treatment in the psoriasis network based on clinical experience for (a) efficacy and (b) safety.

## Appendix Tables

**Appendix Table 1**. League table for efficacy (PASI 90) in lower triangle and safety (serious adverse events (SAE)) in upper triangle. Each cell contains the risk ratio and the respective 95% confidence interval between the treatment in the column and the treatment in the row. Values larger than 1 favour the treatment in the column for both outcomes.

**Appendix Table 2**. League table of probabilities *p*_*i*>*j*_ for efficacy (PASI 90) in lower triangle and safety (serious adverse events (SAE)) in upper triangle. For both outcomes each cell contains the percentage probability that the treatment on the left produces a better outcome than the treatment on the right.

**Appendix Table 3**. Confidence in the evidence for efficacy (PASI 90) as obtained by CINeMA.

**Appendix Table 4**. Probabilities of trusting to use each treatment based on the confidence of the evidence in the respective pairwise comparison.

**Appendix Table 5**. Information on treatment ranking for the psoriasis network in terms of treatment cost. Grey cells correspond to the five top positions.

## Notes

### Competing Interest Statement

The authors have declared no competing interest.

### Funding Statement

No external funding was received

### Author Declarations

All relevant ethical guidelines have been followed and any necessary IRB and/or ethics committee approvals have been obtained.

Any clinical trials involved have been registered with an ICMJE-approved registry such as ClinicalTrials.gov and the trial ID is included in the manuscript.

## References

1. Kibret T, Richer D, Beyene J. Bias in identification of the best treatment in a Bayesian network meta-analysis for binary outcome: a simulation study. Clin Epidemiol. 2014;6:451–460.

2. Trinquart L, Attiche N, Bafeta A, Porcher R, Ravaud P. Uncertainty in Treatment Rankings: Reanalysis of Network Meta-analyses of Randomized Trials. Ann Intern Med. 2016;164(10):666–673.

3. Mbuagbaw L, Rochwerg B, Jaeschke R, et al. Approaches to interpreting and choosing the best treatments in network meta-analyses. Syst Rev. 2017;6:79.

4. Veroniki AA, Straus SE, Rücker G, Tricco AC. Commentary: Is providing uncertainty intervals in treatment ranking helpful in a network meta-analysis? J Clin Epidemiol. 2018;100:122–129

5. Chaimani A, Salanti G, Leucht S, Geddes JR, Cipriani A. Common pitfalls and mistakes in the set-up, analysis and interpretation of results in network meta-analysis: what clinicians should look for in a published article. Evid Based Ment Health. 2017;20(3):88–94.

6. Petropoulou M, Nikolakopoulou A, Veroniki A-A, et al. Bibliographic study showed improving statistical methodology of network meta-analyses published between 1999 and 2015. J Clin Epidemiol. 2017;82:20–28.

7. Chaimani A, Caldwell DM, Li T, Higgins JPT, Salanti G. Additional considerations are required when preparing a protocol for a systematic review with multiple interventions. J Clin Epidemiol. 2017;83:65–74.

8. Salanti G, Ades AE, Ioannidis JP. Graphical methods and numerical summaries for presenting results from multiple-treatment meta-analysis: an overview and tutorial. JClinEpidemiol. 2011;64:163–171.

9. Rücker G, Schwarzer G. Ranking treatments in frequentist network meta-analysis works without resampling methods. BMC Med Res Methodol. 2015;15:58.

10. Chaimani A, Salanti G. Investigating the impact of treatments with scarce evidence in network meta-analysis. 36th ISCB Annual Conference, Utrecht, 2015.

11. Salanti G, Del Giovane C, Chaimani A, Caldwell DM, Higgins JPT. Evaluating the Quality of Evidence from a Network Meta-Analysis. PLoS ONE. 2014;9(7):e99682.

12. Chaimani A, Higgins JPT, Mavridis D, Spyridonos P, Salanti G. Graphical tools for network meta-analysis in STATA. PLoSOne. 2013;8(10):e76654.

13. Brin S, Page L. The Anatomy of a Large-Scale Hypertextual Web Search Engine. Comput Netw. 1998;30:107–117.

14. Blanchet J, Gallego G, Goyal V. A Markov Chain Approximation to Choice Modeling. Oper Res. 2016;64(4):886–905.

15. Sbidian E, Chaimani A, Garcia-Doval I, et al. Systemic pharmacological treatments for chronic plaque psoriasis: a network meta-analysis. Cochrane Database Syst Rev. 2017, 22;12:CD011535.

16. Ibe OC. 4 - Discrete-Time Markov Chains. In: Ibe OC, editor. Markov Process Stoch Model Second Ed. Oxford: Elsevier; 2013. 59–84 p.

17. Grinstead CM, Snell JL. Introduction to Probability. American Mathematical Soc.; 1997.

18. N. Langville A, D. Meyer C. Deeper Inside PageRank. Internet Math. 2004;1.

19. Hutton B, Salanti G, Caldwell DM, et al. The PRISMA Extension Statement for Reporting of Systematic Reviews Incorporating Network Meta-Analyses of Healthcare Interventions: Checklist and Explanations. Ann Intern Med. 2015; 162(11):777-84

20. Puhan MA, Schünemann HJ, Murad MH, et al. A GRADE Working Group approach for rating the quality of treatment effect estimates from network meta-analysis. BMJ. 2014;349:g5630.

21. CINeMA: Confidence in Network Meta-Analysis [Software]. Institute of Social and Preventive Medicine, University of Bern; 2017. Available from: http://cinema.ispm.ch

22. Nikolakopoulou A, Higgins JP, Papakonstantinou T, et al. Assessing Confidence in the Results of Network Meta-Analysis (Cinema). bioRxiv. 2019 Apr 5;597047.

23. Garthwaite PH, Kadane JB, O’Hagan A. Statistical Methods for Eliciting Probability Distributions. J Am Stat Assoc. 2005;100(470):680–700.

24. Morris DE, Oakley JE, Crowe JA. A web-based tool for eliciting probability distributions from experts. Environ Model Softw. 2014;52:1–4.

25. Page L, Brin S, Motwani R, Winograd T. The PageRank Citation Ranking: Bringing Order to the Web. 7th International World Wide Web Conference, p161-172. Brisbane, Australia. 1998.

26. White IR. Network meta-analysis. Stata J. 2015;15(4):951–985.

27. Chaimani A, Salanti G. Visualizing assumptions and results in network meta-analysis: The network graphs package. Stata J. 2015;15(4):905–950.

28. Créquit P, Trinquart L, Yavchitz A, Ravaud P. Wasted research when systematic reviews fail to provide a complete and up-to-date evidence synthesis: the example of lung cancer. BMC Med. 2016;14:8.

29. Créquit P, Trinquart L, Ravaud P. Live cumulative network meta-analysis: protocol for second-line treatments in advanced non-small-cell lung cancer with wild-type or unknown status for epidermal growth factor receptor. BMJ Open. 2016;6(8):e011841.

30. Nikolakopoulou A, Mavridis D, Furukawa TA, et al. Living network meta-analysis compared with pairwise meta-analysis in comparative effectiveness research: empirical study. BMJ. 2018;360:k585.

31. Nikolakopoulou A, Mavridis D, Egger M, Salanti G. Continuously updated network meta-analysis and statistical monitoring for timely decision-making. Stat Methods Med Res. 2018;27(5):1312–1330.

32. Elliott JH, Synnot A, Turner T, et al. Living systematic review: 1. Introduction—the why, what, when, and how. J Clin Epidemiol. 2017;91:23–30.

33. StataCorp. Stata Statistical Software. College Station, TX: StataCorp LP; 2013.

